# Effects of heatwrap and exercise in acute low back pain: a multi-arm randomised controlled trial

**DOI:** 10.1101/2025.09.07.25335260

**Authors:** Claudia Côté-Picard, Jean Tittley, Catherine Mailloux, Amélie Desgagnés, Kadija Perreault, Catherine Mercier, Clermont E. Dionne, Jean-Sébastien Roy, Hugo Massé-Alarie

## Abstract

**Question:** Is a heatwrap combined with exercise more effective in improving symptoms and functional outcomes on short- mid- and long-term than either a heatwrap alone or a sham heatwrap in acute low back pain?

**Design:** A multi-arm randomised controlled trial.

**Participants:** Adults aged 18 to 65 years with a new episode of low back pain (defined as pain for six weeks or less with no low back pain in the three months preceding the current episode).

**Intervention:** Participants were randomly assigned to one of three groups: heatwrap plus exercise (n=34), heatwrap alone (n=33) or sham heatwrap (n=32). All participants were asked to wear the heatwrap eight hours per day for seven days. The exercise group had to perform exercises 30 minutes per day for five days. All participants received evidence-based advice for the management of their pain.

**Outcome measures:** Our primary outcome was pain-related disability at one week measured with the Oswestry Disability Index. Our secondary outcomes were pain intensity, pain-related fear of movement, self-efficacy, pain catastrophizing and perception of change. Outcomes were measured at baseline and at 1, 4, 12, 26 and 52 weeks.

**Data analysis:** We used linear mixed models (LMMs) to compare intervention efficacy on all outcomes at all time-points with intention-to-treat analyses, testing the Group x Time interaction.

**Results:** The analysis included 99 participants. The results for pain-related disability showed no Group x Time interaction (F=0.467; p=0.910), but a statistically significant time effect was observed (F=39.843; p<0.001) as scores decreased for all groups at one week, with an estimated mean difference of −7.8 (95% CI −9.7 to −5.9). All outcomes showed a time effect at all time-points.

**Conclusion:** A combination of heatwrap and exercise did not provide a superior effect on symptoms and functional outcomes on short-, mid and long-term when compared to a heatwrap alone and a sham heatwrap in adults with acute low back pain. Future guidelines should consider these results to formulate recommendations for acute LBP.

**Trial registration:** ClinicalTrials.gov Identifier: NCT03986047

## Introduction

Low back pain (LBP) remains the principal cause of years lived with disability globally, as confirmed by the 2021 Global Burden of Disease study.^1^ Acute LBP, defined as pain for less than six weeks, significantly improves in the first six weeks after onset.^2, 3^ Two-thirds of patients with acute or subacute LBP (pain for less than 12 weeks), however, continue to experience pain after one year.^4^ LBP that is not explained by a condition beyond the lumbar spine (e.g. kidney), a neurological impairment, a serious pathology (e.g. fracture, infection), or an inflammatory disease (e.g. arthritis) is considered as being non-specific.^5^ First-line care recommended for patients with non-specific LBP consists of minimal intervention, including reassurance about the generally benign nature of LBP, advice to remain active and self-manage with non-pharmacological strategies.^5, 6^

Interventions recommended specifically in acute LBP include superficial heat with a moderate quality of evidence, and massage, spinal manipulation and acupuncture with a low quality of evidence.^5, 7^ High-quality systematic reviews and meta-analyses showed that heatwrap had a small to moderate short-term effect on pain relief in acute and subacute LBP compared to an oral placebo, with a very low to moderate certainty of evidence.^8, 9^ Exercises are endorsed by most clinical practice guidelines in chronic LBP, but discrepancies exist regarding their usefulness in acute LBP.^7, 10, 11^ Recent systematic reviews and meta-analyses showed that exercise was not effective in acute LBP compared to a placebo, no treatment or other conservative treatments (e.g. manipulations, advice, usual care), with a very low to moderate certainty of evidence.^9, 12^ In contrast, a 2021 network meta-analysis of acute and subacute LBP treatments showed exercise to be effective in decreasing pain and disability in the short-term compared to sham or placebo treatments and no treatment.^13^ A combination of heatwrap and exercise was also shown to provide a small short-term effect on pain relief when compared to either the heatwrap or exercise alone, or to a booklet, with a moderate certainty of evidence.^8^ Important limitations of prior randomised controlled trials (RCTs) on heatwrap in acute LBP are the lack of independent research, as all trials were funded by heatwrap manufacturers, and the fact that none assessed its long-term effect.^8, 9, 14–21^

High-quality placebo- or sham-controlled trials are needed to strengthen recommendations on heat therapy and exercise in acute LBP. As heatwrap was shown to provide pain relief in acute LBP, it could potentiate the effect of exercise in the acute phase, and the combination of these interventions might contribute to reduce disability and pain in the short-term but also to prevent the transition from acute to chronic LBP. The aim of this RCT was to compare the effect of a heatwrap used alone or combined with exercise to a sham heatwrap in a one-week intervention on symptoms and functional outcomes at short-, mid- and long-term in adults with acute LBP. We hypothesised that the combination of heatwrap and exercise would have a positive effect on all outcomes on short-, mid- and long-term compared to both the heatwrap alone and the sham heatwrap.

## Material and methods

### Design

This was a multi-arm parallel RCT trial with a 1:1:1 allocation ratio, at a single research center in Québec City, Canada. The study received ethical approval from the Research Ethics Committee of the *CIUSSS de la Capitale-Nationale* (#2019-1731, RIS – CER CIUSSS-CN) and follows the Consolidated Standards of Reporting Trials (CONSORT) and the multi-arm extension.^22, 23^ The protocol was published previously,^24^ and registered on Clinicaltrials.gov (NCT03986047).

Participants provided their written consent before enrolling in the study. There were two visits to the research center over one week (W_0_ and W_1_), and follow-ups at 4, 12, 26 and 52 weeks (W_4_, W_12_, W_26_ and W_52_). At W_0_, questionnaires on sociodemographic (Canadian Minimum Dataset for low back pain – CMDS)^25^ and prognostic characteristics (STarT Back Screening Tool),^26^ and patient reported outcome measurements (PROMs) were self-administered by participants. Then, participants were randomly allocated to one of three intervention groups: heatwrap and exercise, heatwrap alone, or sham heatwrap, and met a physical therapist blinded from the baseline assessment who gave them instructions on their attributed intervention in a 60-minute meeting. Participants were asked to perform the intervention at home for 7 days, then to complete the PROMs online at W_1_, W_4_, W_12_, W_26_ and W_52_. A second visit to the research center took place after the intervention week (W_1_) in which participants could ask questions about their condition and final general advice was given.

### Participants

The recruitment took place from April 2019 to March 2023 via email listings to the Université Laval’s community, social media ads, and through the Quebec Back Pain Consortium database.^27^ We recruited adults aged 18 to 65 years with acute non-specific LBP (pain for 6 weeks or less with or without leg pain)^28^ who understood written French, who had a score ≥ 10 on the Oswestry Disability Index (ODI) and who experienced limited daily life activities for more than one day. The ODI cut-off was chosen so that we could measure a minimal clinically important difference (MCID), that is 9 points in patients with acute LBP.^29^ We excluded those who had LBP in the three months preceding the current episode, signs of a serious pathology or peripheral/central neural impairment, who were pregnant, diagnosed with rheumatoid arthritis, fibromyalgia, had important skin lesion in the lumbar area, or cognitive impairments. We also excluded those who had a history of spinal surgery, a recent change in medication that could influence pain (antidepressants, opioids, etc.), or those who had a corticosteroid injection in the 6 weeks prior to their enrollment in the study.

### Randomisation and blinding

An independent research assistant generated a randomisation list with a computer random number generator in random blocks of 3 or 6 participants. The allocation was concealed in sealed and opaque envelopes. Stratification was done by sex and ODI score (≤19: low or ≥20: moderate disability) ODI dichotomisation was based on Fairbank et al (1980).^30^ Participants were unaware of the nature of interventions in the other groups.

### Interventions

1. Heatwrap and exercise: Participants were asked to wear a heatwrap (ThermaCare^®^ Back Pain Therapy HeatWraps, Bridges Consumer Healthcare, TN, USA) to the lower lumbar spine area during 8 hours per day for 7 days, and to realise their assigned exercise program during 30 minutes per day for 5 days. The heatwrap heats up to 40°C in 30 minutes, and this temperature is maintained for 8 hours. It is for single use only, so 7 were distributed to participants. No standardised application parameters are recommended for heatwrap, so our parameters were based on other studies that used it for 8 hours per day for 1 to 14 days. ^14–21^ The exercise program was prescribed by the physical therapist and individualised to each participant according to the findings of a physical assessment and included 3 or 4 exercises from a predetermined list of four different categories: functional task (e.g. sitting to standing, lifting), trunk muscle activation, lumbar spine mobility, and preferential direction if applicable. The Consensus on Exercise Reporting Template (CERT) and the complete exercise list and description were published as Additional files in the study protocol.^24^
2. Heatwrap alone: Participants were asked to wear the same heatwrap with the same parameters as the “Heatwrap and exercise” group.
3. Sham heatwrap: Participants were asked to wear a heatwrap that was previously cooled down to the room temperature, in a same manner as in the other groups to control for the potential supportive and sensory influence of the heatwrap. The heatwrap bag was previously opened so it was not warm when distributed to participants.

In addition, all participants received advice on acute LBP management. It included reassurance about the generally favorable prognosis of acute LBP, encouragement to remain active and continue to work or return to work as soon as possible, and to avoid bed rest.^24^ Participants were asked to complete a logbook everyday during the intervention week to record their adherence to interventions and any adverse event. They were instructed to avoid seeking treatment by other healthcare professionals for their LBP during the intervention week if possible.

At the second visit to the research center (W_1_), after the intervention week, three general trunk muscle activation exercises were given to participants in the heatwrap alone and the sham heatwrap groups. These exercises were adapted to each participant’s condition in the same manner as for the heatwrap and exercise group.^24^ This decision was taken to offer an active intervention to all patients (especially the sham group) which is in line with the recommendations of encouraging physical activity.

### Outcome measures

Our primary outcome was pain-related disability at W_1_ measured with ODI. This is a self-administered questionnaire with 10 questions to answer on a 6-point Likert scale, where 0 means no disability and 5 means maximal pain-related disability (MCID = 9).^29, 31^ Secondary outcomes were pain-related disability at other time-points than W_1_, the average pain intensity over the last week (numerical pain rating scale – NPRS),^32^ pain-related fear of movement (17-item version Tampa Scale of Kinesiophobia – TSK-17),^33^ self-efficacy (short version of the Chronic Pain Self-Efficacy Scale – CPSES-6),^34^ perception of change (15-point global rating of change – GroC),^35^ pain catastrophizing, measured with the Pain Catastrophizing Scale (PCS),^36^ and the average pain intensity measured each day of the intervention week with the NPRS. ODI, NPRS (1-week average), TSK-17, CPSES-6, and PCS were all self-administered online at W_0_, W_1_, W_4_, W_12_, W_26_ and W_52_, and GRoC at W_1_, W_4_ W_12_, W_26_ and W_52_. The percentage of participants using interventions for their LBP (exercises, opioid, corticosteroid injection, and psychological support) were extracted from the CMDS at W_0_, W_4_, W_12_, W_26_ and W_52_. All outcomes were measured online via LimeSurvey, except for the average pain intensity recorded each day of the intervention week in participants’ logbook (day 1 through 7 – D_1_, D_2_, D_3_, D_4_, D_5_, D_6_ and D_7_).

### Sample size

We calculated our sample size using G*Power v3.1.9.7 for a between-group difference in our primary outcome at 1 week with an effect size of 0.8, an alpha level of 0.05, a power of 0.8, and a 10% loss at follow-up, resulting in 23 participants per group. We based our effect size on studies that assessed the effect of heatwrap in acute LBP.^14, 18–21^ A network meta-analysis from Gianola et al. (2021) pooled their effect sizes and obtained standardised mean differences (SMD) of −1.38 for pain reduction and −0.59 for disability at 1 week in favor of heatwrap when compared to inert treatment.^13^ Over the course of the trial, additional funding allowed to recruit ten more participants per group, which increased the power to 0.94 when calculated *a posteriori*.

### Data analysis

We used linear mixed models (LMMs) to compare intervention efficacy on all outcomes with intention-to-treat analyses and a random effect for participants using IBM SPSS statistics v.30.0. We verified normality and variance homogeneity by examining the residuals’ distribution, skewness and kurtosis. We compared the effect of all three interventions on pain-related disability, average pain intensity over the last week, pain-related fear of movement, self-efficacy and catastrophizing at 6 time-points, (W_0_, W_1_, W_4_, W_12_, W_26_, W_52_), perception of change at 5 time-points (W_1_, W_4_, W_12_, W_26_ and W_52_), and average pain intensity each day of the intervention week at 7 time-points (D_1_, D_2_, D_3_, D_4_, D_5_, D_6_ and D_7_) testing the Group x Time interaction. As recommended, we included our stratification variables (sex and ODI [≤19 and ≥20]), and a prognostic factor (age)^37^ as covariates in the models to increase their statistical power.^38, 39^ We did not adjust our alpha level (0.05) for multiplicity as we predetermined a primary outcome (ODI at 1 week),^40^ and we treated the analyses of secondary outcomes as exploratory. If our models obtained a p<0.05 for the Group x Time interaction, we used post-hoc analyses to find where the differences were. Our models use a restricted maximum likelihood estimation approach, that assumes data to be missing at random, allowing all available data to be included in the models.^41^.

### Sensitivity analyses

We performed a sensitivity analysis with a per-protocol approach for all outcomes by including only participants who had worn the wrap (heated or unheated) more than 75% of the time they were asked to wear it (average of six hours and more per day), and those in the heatwrap and exercise group who performed exercises for at least three days out of five. We also performed a sensitivity analysis for our primary outcome (ODI) using the same model as our primary analysis, but in including the proportion of participants who were performing exercises across the study as a covariate.

### Deviations from protocol

We specified in the protocol that step counts recorded with a fit watch during the intervention week as a proxy of physical activity would be a secondary outcome, but we had technical issues with the equipment and obtained unreliable data, so it was excluded from the analysis. We used the 17-item TSK instead of the 11-item version as it was shown to be more valid and reliable.^33^ Pain catastrophizing, measured with the Pain Catastrophizing Scale (PCS) at W_0_, W_1_, W_4_, W_12_, W_26_ and W_52_,^36^ and the average pain intensity measured each day of the intervention week with the NPRS were added as secondary outcomes. We initially reported in the protocol^24^ that the last follow-up would be at 26 weeks, but additional funding allowed to add a follow-up at 52 weeks. Additional funding also allowed to recruit 10 more participants per group. We decided to use linear mixed models instead of generalised linear mixed models as our data respected normality and variance homogeneity.

## Results

### Participants

From April 2019 to March 2023, 100 individuals were recruited in the trial out of 315 who initially communicated their interest. One participant from the heatwrap and exercise group was excluded after randomisation because he received a diagnosis of a lumbar vertebrae fracture that likely occurred at the same moment as the beginning of the LBP episode. Baseline participants’ demographic and clinical characteristics are presented in Table 1. Most participants were female (62%), the mean (standard deviation [SD]) age was 36.4 (12.8) years, and 63% had completed a bachelor’s degree or higher education. The mean (SD) duration of the current LBP episode was 25.2 (11.9) days, and the mean disability and pain intensity scores at baseline were 22.8% (10.9) and 5.1/10 (1.9), respectively. The mean (SD) duration that participants self-declared to have worn their attributed wrap each day of the intervention week was 7.5 (1.1) hours for the heatwrap and exercise group, 8.2 (1.8) hours for the heatwrap alone group, and 8.0 (1.7) hours for the sham wrap group. The participants in the heatwrap plus exercise group self-declared to have performed exercises on an average of 6.2 days during the intervention week. Only 2 participants did not return their logbook as their second visit to the research center was cancelled (Covid-19 lockdown and illness). Of those who returned their logbook, all performed at least 3 days of exercises. One participant in the heatwrap alone group only completed 3 intervention days. Overall, 97 participants (97%) provided data for the primary outcome at W_1_, 94 (94%) at W_4_, 89 (89%) at W_12_, 87 (87%) at W_26_, and 84 (84%) at W_52_ (Figure 1).

**Figure 1.**
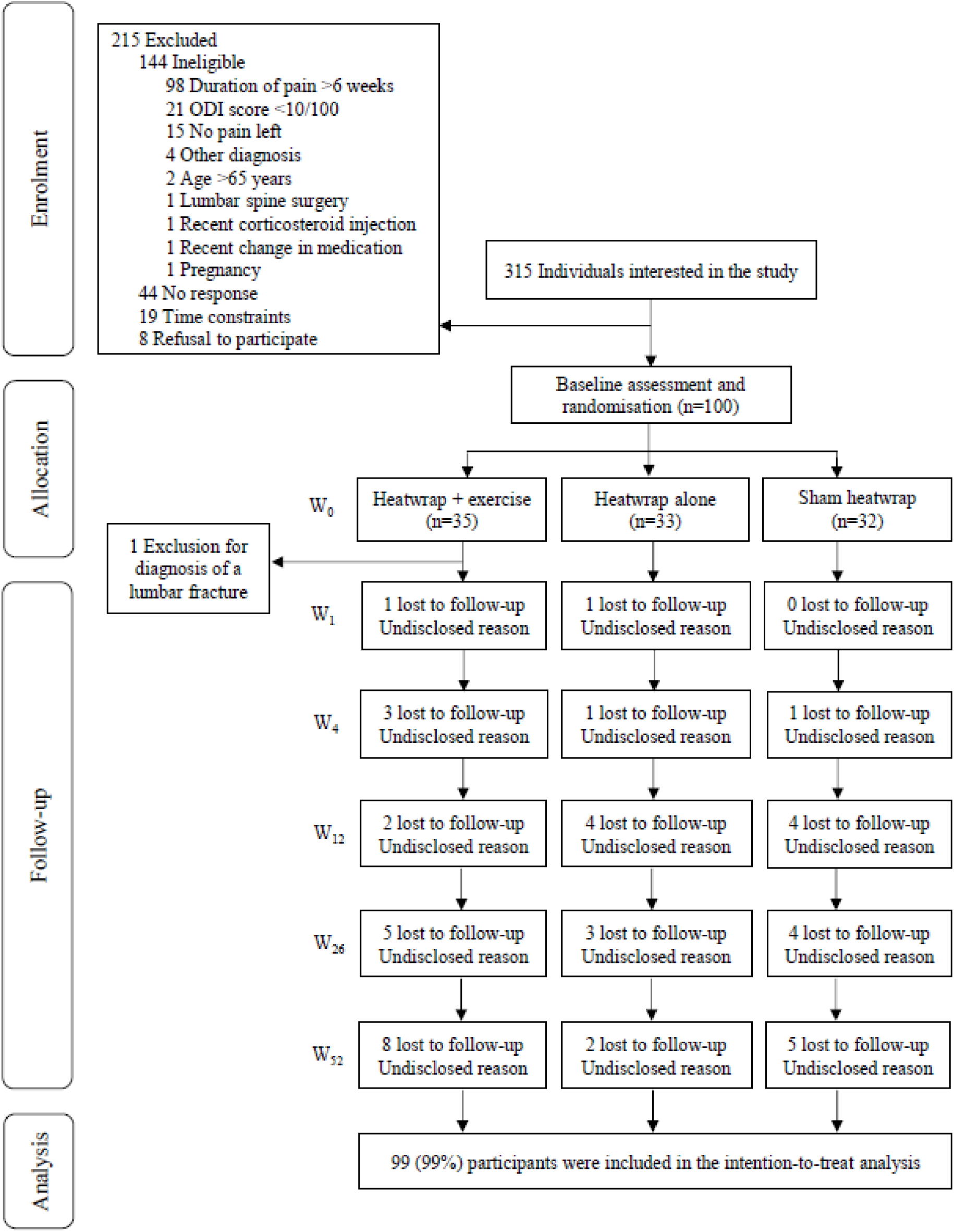
Trial flowchart

**Table 1.** Baseline demographic and clinical characteristics of study participants.

|  | Heatwrap and exercise (n=34) | Heatwrap alone (n=33) | Sham heatwrap (n=32) | Total (n=99) |
| --- | --- | --- | --- | --- |
| **Demographic characteristics** |  |  |  |  |
| Sex, n of females (%) | 20 (59) | 21 (64) | 20 (63) | 61 (62) |
| Age (years), mean [SD] | 35.1 [12.1] | 36.9 [13.6] | 37.3 [13.0] | 36.4 [12.8] |
| BMI (kg/m<sup>2</sup>), mean [SD] | 25.3 [3.7] | 27.1 [5.5] | 27.6 [5.8] | 26.7 [5.1] |
| Employment status, n (%) |  |  |  |  |
| Working full time | 16 (47) | 11 (33) | 13 (41) | 40 (40) |
| Working part time | 7 (21) | 14 (42) | 10 (31) | 31 (31) |
| Unemployed/ looking for a job | 3 (9) | 2 (6) | 1 (3) | 6 (6) |
| Sick leave/ maternity leave | 1 (3) | 1 (3) | 0 (0) | 2 (2) |
| Disabled due to back pain | 0 (0) | 0 (0) | 1 (3) | 1 (1) |
| Disabled for other reasons | 0 (0) | 1 (3) | 1 (3) | 2 (2) |
| Student | 16 (47) | 14 (42) | 16 (50) | 46 (47) |
| Temporary layoff | 2 (6) | 1 (3) | 0 (0) | 3 (3) |
| Retired | 0 (0) | 1 (3) | 2 (6) | 3 (3) |
| Unknown | 0 (0) | 1 (3) | 0 (0) | 1 (1) |
| Highest education level completed, n (%) |  |  |  |  |
| High school graduate | 2 (6) | 4 (12) | 1 (3) | 7 (7) |
| Certificate or Diploma | 11 (32) | 8 (24) | 11 (34) | 30 (30) |
| Bachelor's degree or higher | 21 (62) | 21 (64) | 20 (63) | 62 (63) |
| **Clinical characteristics** |  |  |  |  |
| Participants with a history of LBP, n (%) | 24 (71) | 20 (61) | 19 (59) | 63 (64) |
| LBP episode duration (days), mean [SD] | 24.2 [12.2] | 26.2 [11.2] | 25.4 [12.4] | 25.2 [11.9] |
| ODI, mean [SD] | 22.9 [10.6] | 23.2 [10.5] | 22.4 [11.9] | 22.8 [10.9] |
| NPRS*, mean [SD] | 4.7 [1.9] | 5.5 [1.9] | 5.2 [1.7] | 5.1 [1.9] |
| TSK-11, mean [SD] | 36.7 [7.4] | 35.6 [6.3] | 36.7 [6.9] | 36.3 [6.8] |
| SBST, n (%) |  |  |  |  |
| Low risk | 20 (59) | 19 (58) | 17 (53) | 56 (57) |
| Medium risk | 13 (38) | 12 (36) | 12 (38) | 37 (37) |
| High risk | 1 (3) | 2 (6) | 3 (9) | 6 (6) |
| n, number of participants; LBP, low back pain; 95% CI, 95% confidence interval; BMI, body mass index; NPRS, Numerical Pain Rating Scale; ODI, Oswestry Disability Index; TSK-11, Tampa Scale for Kinesiophobia; SBST, Start back screening tool.<br>*The NPRS score is the average pain intensity over the last week. |  |  |  |  |

### Primary outcome

The primary analysis for pain-related disability showed no significant Group x Time interaction (F=0.467; p=0.910), but a statistically significant time effect was observed (F=39.843; p<0.001) as ODI scores decreased for all groups at W_1_, with an estimated mean difference (MD) of −7.8 (95% CI −9.7 to −5.9) (Figure 2A; Table 2). At W_1_, ODI scores for the heatwrap and exercise group were not significantly different from the heatwrap alone (MD −1.6, 95% CI −6.2 to 3.0), or the unheated wrap (MD −0.5, 95% CI −5.0 to 4.2) groups. Within- and between-group estimated MDs for all outcomes and all time-points are presented in Table 2 and Table 3, respectively.

**Figure 2.**
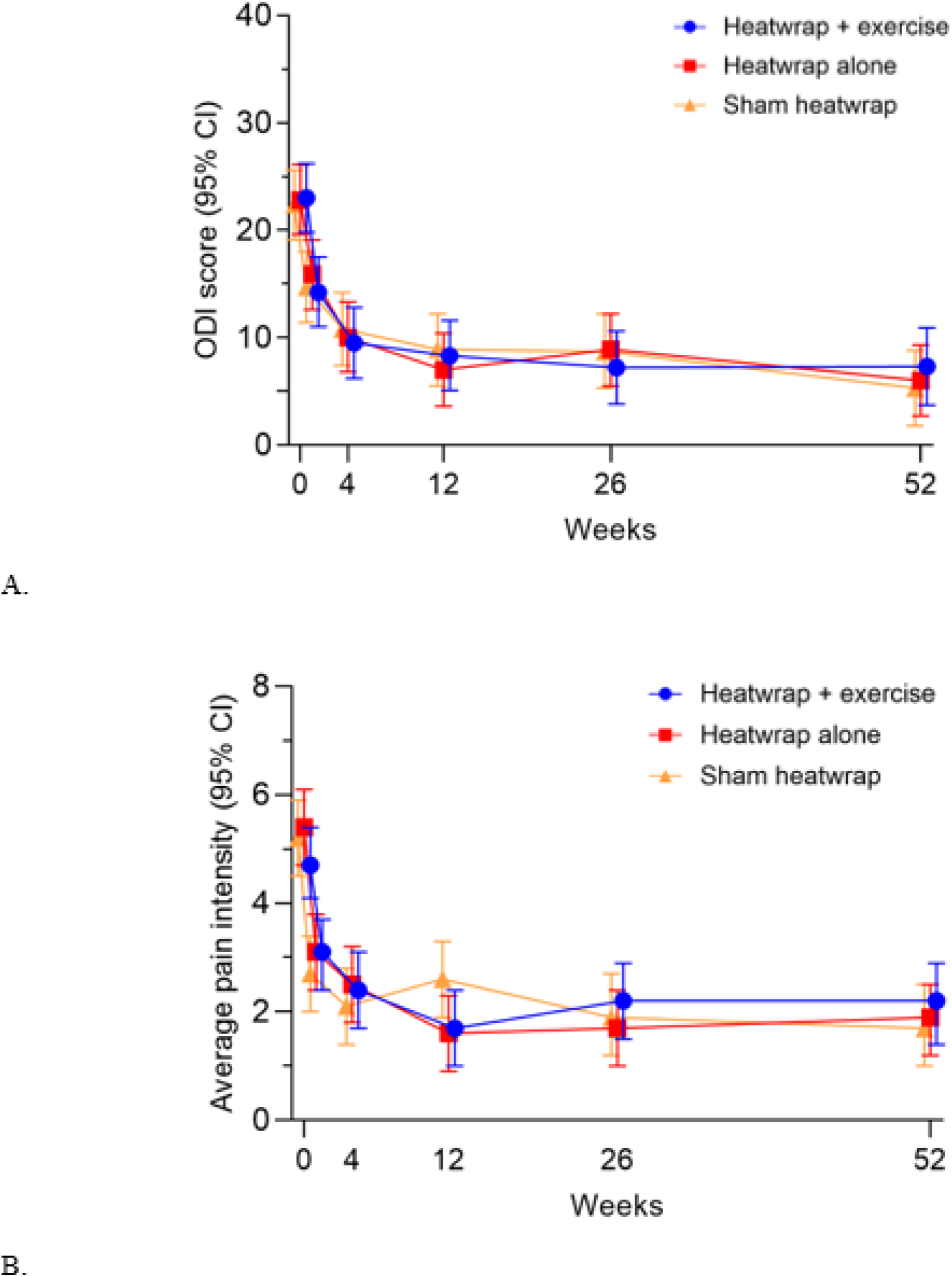
Main effect of time in A. Pain-related disability levels (ODI scores) and B. Average pain intensity and 95% CI.

**Table 2.** Within-group estimated mean differences at each time-point for each outcome and 95% confidence intervals (95% CI)

|  | **Heatwrap and exercise** | **Heatwrap alone** | **Sham heatwrap** | **Overall – Time effect** |
| --- | --- | --- | --- | --- |
| **ΔODI (0 to 100) with W<sub>0</sub>** |  |  |  |  |
| W<sub>1</sub> | -8.8 (-12.0 to -5.5) | -7.0 (-10.2 to -3.7) | -7.7 (-11.0 to -4.4) | -7.8* (-9.7 to -5.9) |
| W<sub>4</sub> | -13.5 (-17.4 to -9.6) | -12.8 (-16.8 to -8.9) | -11.6 (-15.6 to -7.5) | -12.6* (-14.9 to -10.3) |
| W<sub>12</sub> | -14.7 (-18.8 to -10.5) | -15.8 (-20.1 to -11.5) | -13.5 (-17.8 to -9.2) | -14.7* (-17.1 to -12.2) |
| W<sub>26</sub> | -15.8 (-20.2 to -11.4) | -14.0 (-18.4 to -9.6) | -13.6 (-18.1 to -9.1) | -14.5* (-17.0 to -11.9) |
| W<sub>52</sub> | -15.7 (-20.3 to -11.1) | -16.8 (-21.3 to -12.4) | -17.1 (-21.7 to -12.5) | -16.5* (-19.2 to -13.9) |
| **ΔNPRS (0 to 10) with W<sub>0</sub> – Average pain intensity in the last 7 days** |  |  |  |  |
| W<sub>1</sub> | -1.7 (-2.4 to -1.0) | -2.4 (-3.1 to -1.6) | -2.4 (-3.2 to -1.7) | -2.2* (-2.5 to -1.7) |
| W<sub>4</sub> | -2.3 (-3.1 to -1.5) | -3.0 (-3.8 to -2.1) | -3.1 (-3.9 to -2.2) | -2.8* (-3.3 to -2.3) |
| W<sub>12</sub> | -3.0 (-3.9 to -2.2) | -3.8 (-4.7 to -3.0) | -2.5 (-3.4 to -1.7) | -3.1* (-3.6 to -2.6) |
| W<sub>26</sub> | -2.6 (-3.4 to -1.7) | -3.7 (-4.6 to -2.9) | -3.2 (-4.1 to -2.3) | -3.2* (-3.7 to -2.7) |
| W<sub>52</sub> | -2.6 (-3.5 to -1.7) | -3.6 (-4.4 to -2.7) | -3.5 (-4.4 to -2.5) | -3.2* (-3.7 to -2.7) |
| **ΔTSK-17 (17 to 68) with W<sub>0</sub>** |  |  |  |  |
| W<sub>1</sub> | -3.8 (-5.7 to -1.8) | -2.2 (-4.2 to -0.3) | -3.4 (-5.4 to -1.5) | -3.1* (-4.3 to -2.0) |
| W<sub>4</sub> | -5.2 (-7.3 to -3.1) | -4.0 (-6.1 to -1.9) | -4.5 (-6.7 to -2.3) | -4.6* (-5.8 to -3.3) |
| W<sub>12</sub> | -6.0 (-8.2 to -3.9) | -3.3 (-5.5 to -1.1) | -5.4 (-7.6 to -3.2) | -4.9* (-6.2 to -3.7) |
| W<sub>26</sub> | -4.2 (-6.4 to -2.0) | -4.0 (-6.2 to -1.8) | -4.6 (-6.8 to -2.3) | -4.3* (-5.5 to -3.0) |
| W<sub>52</sub> | -5.2 (-7.5 to -2.9) | -4.8 (-7.0 to -2.6) | -5.3 (-7.6 to -2.9) | -5.1* (-6.4 to -3.8) |
| **ΔCPSES-6 (6 to 60) with W<sub>0</sub>** |  |  |  |  |
| W<sub>1</sub> | 0.5 (-0.1 to 1.0) | 0.5 (-0.1 to 1.0) | 0.2 (-0.3 to 0.8) | 0.4<sup>□</sup> (0.1 to 0.7) |
| W<sub>4</sub> | 0.9 (0.3 to 1.5) | 0.7 (0.1 to 1.3) | 0.6 (-0.0 to 1.3) | 0.7* (0.4 to 1.1) |
| W<sub>12</sub> | 0.9 (0.3 to 1.6) | 0.7 (0.1 to 1.4) | 0.7 (-0.0 to 1.4) | 0.8* (0.4 to 1.2) |
| W<sub>26</sub> | 0.7 (0.0 to 1.4) | 0.8 (0.1 to 1.5) | 0.3 (-0.4 to 1.0) | 0.6<sup>□</sup> (0.2 to 1.0) |
| W<sub>52</sub> | 0.7 (-0.1 to 1.4) | 1.0 (0.3 to 1.8) | 0.8 (0.1 to 1.5) | 0.8* (0.4 to 1.3) |
| **ΔPCS (0 to 52) with W<sub>0</sub>** |  |  |  |  |
| W<sub>1</sub> | -4.2 (-6.9 to -1.5) | -5.6 (-8.3 to -2.9) | -5.7 (-8.4 to -2.9) | -5.2* (-6.7 to -3.6) |
| W<sub>4</sub> | -8.5 (-11.6 to -5.4) | -7.9 (-11.0 to -4.8) | -6.7 (-9.8 to -3.5) | -7.7* (-9.5 to -5.9) |
| W<sub>12</sub> | -9.1 (-12.3 to -5.9) | -8.5 (-11.7 to -5.2) | -8.1 (-11.3 to -4.8) | -8.5* (-10.4 to -6.7) |
| W<sub>26</sub> | -9.2 (-12.5 to -5.9) | -9.2 (-12.4 to -6.0) | -8.7 (-12.0 to -5.4) | -9.1* (-10.9 to -7.2) |
| W<sub>52</sub> | -6.5 (-9.9 to -3.1) | -7.7 (-11.0 to -4.5) | -9.3 (-12.8 to -5.9) | -7.8* (-9.8 to -5.9) |
| **ΔGroC (-7 to 7) with W<sub>0</sub>** |  |  |  |  |
| W<sub>4</sub> | 0.3 (-0.9 to 1.6) | 2.4 (1.1 to 3.7) | 0.5 (-0.9 to 1.8) | 1.1<sup>□</sup> (0.3 to 1.8) |
| W<sub>12</sub> | 0.5 (-0.9 to 1.9) | 3.1 (1.7 to 4.6) | -0.3 (-1.7 to 1.1) | 1.1<sup>□</sup> (0.3 to 1.9) |
| W<sub>26</sub> | 0.2 (-1.3 to 1.6) | 3.5 (2.1 to 5.0) | 0.4 (-1.0 to 1.9) | 1.4<sup>□</sup> (0.5 to 2.2) |
| W<sub>52</sub> | 0.3 (-1.2 to 1.8) | 3.0 (1.6 to 4.5) | 0.2 (-1.3 to 1.8) | 1.2<sup>□</sup> (0.3 to 2.0) |
| **ΔNPRS (0 to 10) – Average daily pain intensity with D<sub>1</sub>** |  |  |  |  |
| D<sub>2</sub> | -0.5 (-0.9 to -0.1) | -0.4 (-0.8 to -0.0) | -0.6 (-1.0 to -0.2) | -0.5 (-0.8 to -0.3) |
| D<sub>3</sub> | -0.9 (-1.4 to -0.4) | -0.7 (-1.2 to -0.2) | -0.8 (-1.3 to -0.3) | -0.8 (-1.1 to -0.5) |
| D<sub>4</sub> | -1.0 (-1.5 to -0.5) | -0.8 (-1.3 to -0.2) | -0.6 (-1.1 to -0.0) | -0.8 (-1.1 to -0.5) |
| D<sub>5</sub> | -1.1 (-1.7 to -0.5) | -0.6 (-1.2 to -0.1) | -0.7 (-1.3 to -0.2) | -0.8 (-1.1 to -0.5) |
| D<sub>6</sub> | -1.2 (-1.8 to -0.6) | -0.8 (-1.4 to -0.3) | -0.5 (-1.1 to 0.0) | -0.9 (-1.2 to -0.5) |

|  | **Heatwrap and exercise** | **Heatwrap alone** | **Sham heatwrap** | Overall – Time effect |
| --- | --- | --- | --- | --- |
| D<sub>7</sub> | -1.7 (-2.3 to -1.1) | -1.2 (-1.8 to -0.6) | -0.8 (-1.3 to -0.2) | -1.2 (-1.6 to -0.9) |
| ODI, Oswestry Disability Index; NPRS, Numerical Pain Rating Scale; TSK-17, 17-item Tampa Scale of Kinesiophobia; CDSES-6, Chronic Disease Self-Efficacy Scale short version; PCS, Pain Catastrophizing Scale; GRoC, 15-point global rating of change; W<sub>1</sub>, week 1; W<sub>4</sub>, week 4; W<sub>12</sub>, week 12; W<sub>26</sub>, week 26; W<sub>52</sub>, week 52; D<sub>1</sub>, day 1; D<sub>2</sub>, day 2; D<sub>3</sub>, day 3; D<sub>4</sub>, day 4; D<sub>5</sub>, day 5; D<sub>6</sub>, day 6; D<sub>7</sub>, day 7.<br>□ p ≤0.01; *p <0.001 |  |  |  |  |

**Table 3.** Between-group estimated mean difference at each time-point for each outcome and 95% confidence intervals (95% CI)

|  | Mean difference (95% confidence interval) with sham heatwrap |  | Mean difference between heatwrap groups (H + E minus H alone) |
| --- | --- | --- | --- |
|  | Heatwrap and exercise | Heatwrap alone |  |
| ΔODI (0 to 100) | | | |
| W<sub>0</sub> | 0.6 (-3.9 to 5.2) | 0.5 (-4.1 to 5.1) | 0.2 (-4.4 to 4.7) |
| W<sub>1</sub> | -0.5 (-5.0 to 4.2) | 1.2 (-3.4 to 5.8) | -1.6 (-6.2 to 3.0) |
| W<sub>4</sub> | -1.3 (-6.0 to 3.5) | -0.8 (-5.5 to 4.0) | -0.5 (-5.1 to 4.1) |
| W<sub>12</sub> | -0.6 (-5.4 to 4.1) | -1.9 (-6.6 to 2.9) | 1.3 (-3.4 to 6.0) |
| W<sub>26</sub> | -1.5 (-6.3 to 3.3) | 0.2 (-4.7 to 5.0) | -1.7 (-6.4 to 3.1) |
| W<sub>52</sub> | 2.0 (-3.0 to 7.0) | 0.7 (-4.1 to 5.6) | 1.3 (-3.6 to 6.2) |
| ΔNPRS (0 to 10) – Average pain intensity in the last 7 days | | | |
| W<sub>0</sub> | -0.4 (-1.4 to 0.5) | 0.3 (-0.7 to 1.2) | -0.7 (-1.7 to 0.3) |
| W<sub>1</sub> | 0.3 (-0.6 to 1.3) | 0.4 (-0.6 to 1.3) | -0.0 (-1.0 to 0.9) |
| W<sub>4</sub> | 0.3 (-0.7 to 1.3) | 0.4 (-0.6 to 1.4) | -0.1 (-1.1 to 0.9) |
| W<sub>12</sub> | -0.9 (-1.9 to 0.1) | -1.0 (-2.0 to -0.0) | 0.1 (-0.8 to 1.1) |
| W<sub>26</sub> | 0.3 (-0.8 to 1.3) | -0.2 (-1.3 to 0.8) | 0.5 (-0.5 to 1.5) |
| W<sub>52</sub> | 0.5 (-0.6 to 1.5) | 0.1 (-0.9 to 1.2) | 0.3 (-0.7 to 1.3) |
| ΔTSK-17 (17 to 68) | | | |
| W<sub>0</sub> | 0.1 (-3.3 to 3.5) | -1.2 (-4.6 to 2.2) | 1.3 (-2.0 to 4.7) |
| W<sub>1</sub> | -0.2 (-3.6 to 3.2) | 0.02 (-3.4 to 3.4) | -0.3 (-3.6 to 3.1) |
| W<sub>4</sub> | -0.6 (-4.1 to 2.8) | -0.7 (-4.1 to 2.7) | 0.1 (-3.3 to 3.5) |
| W<sub>12</sub> | -0.5 (-3.9 to 2.9) | 0.9 (-2.6 to 4.3) | -1.4 (-4.8 to 2.0) |
| W<sub>26</sub> | 0.5 (-3.0 to 3.9) | -0.7 (-4.2 to 2.8) | 1.1 (-2.3 to 4.6) |
| W<sub>52</sub> | 0.2 (-3.4 to 3.8) | -0.7 (-4.3 to 2.8) | 0.9 (-2.6 to 4.4) |
| ΔCPSES-6 (6 to 60) | | | |
| W<sub>0</sub> | -0.3 (-1.0 to 0.5) | -0.2 (-1.0 to 0.5) | -0.0 (-0.8 to 0.7) |
| W<sub>1</sub> | -0.0 (-0.8 to 0.8) | 0.1 (-0.7 to 0.8) | -0.1 (-0.8 to 0.7) |
| W<sub>4</sub> | 0.0 (-0.7 to 0.8) | -0.2 (-0.9 to 0.6) | 0.2 (-0.6 to 0.9) |
| W<sub>12</sub> | 0.0 (-0.7 to 0.8) | -0.1 (-0.9 to 0.6) | 0.2 (-0.6 to 0.9) |
| W<sub>26</sub> | 0.2 (-0.6 to 1.0) | 0.3 (-0.5 to 1.1) | -0.1 (-0.9 to 0.7) |
| W<sub>52</sub> | -0.4 (-1.2 to 0.5) | 0.1 (-0.8 to 0.9) | -0.4 (-1.2 to 0.4) |
| ΔPCS (0 to 52) | | | |
| W<sub>0</sub> | -1.5 (-5.5 to 2.6) | 2.1 (-2.0 to 6.1) | -3.5 (-7.5 to 0.5) |
| W<sub>1</sub> | 0.0 (-4.0 to 4.1) | 2.1 (-2.0 to 6.2) | -2.1 (-6.1 to 1.9) |
| W<sub>4</sub> | -3.3 (-7.4 to 0.9) | 0.9 (-3.3 to 5.0) | -4.1 (-8.2 to -0.1) |
| W<sub>12</sub> | -2.5 (-6.6 to 1.7) | 1.7 (-2.5 to 5.8) | -4.1 (-8.3 to 0.0) |
| W<sub>26</sub> | -2.0 (-6.2 to 2.3) | 1.6 (-2.6 to 5.8) | -3.5 (-7.7 to 0.6) |
| W<sub>52</sub> | 1.4 (-3.0 to 5.8) | 3.6 (-0.7 to 8.0) | -2.3 (-6.6 to 2.0) |
| ΔGroC (-7 to 7) | | | |
| W<sub>1</sub> | 0.1 (-1.5 to 1.7) | -2.5* (-4.1 to -0.9) | 2.6* (1.0 to 4.1) |
| W<sub>4</sub> | -0.0 (-1.6 to 1.6) | -0.6 (-2.2 to 1.0) | 0.6 (-1.0 to 2.1) |
| W<sub>12</sub> | 0.9 (-0.7 to 2.5) | 1.0 (-0.6 to 2.5) | -0.1 (-1.7 to 1.5) |
| W<sub>26</sub> | -0.2 (-1.8 to 1.5) | 0.6 (-1.0 to 2.2) | -0.8 (-2.4 to 0.8) |
| W<sub>52</sub> | 0.2 (-1.5 to 1.9) | 0.3 (-1.3 to 2.0) | -0.1 (-1.8 to 1.5) |
| ΔNPRS (0 to 10) – Average daily pain intensity | | | |
| D<sub>1</sub> | 0.7 (-0.2 to 1.7) | 0.3 (-0.6 to 1.2) | 0.5 (-0.4 to 1.4) |
| D<sub>2</sub> | 0.8 (-0.1 to 1.7) | 0.5 (-0.4 to 1.4) | 0.4 (-0.5 to 1.3) |
| D<sub>3</sub> | 0.6 (-0.3 to 1.5) | 0.3 (-0.6 to 1.2) | 0.3 (-0.6 to 1.2) |
| D<sub>4</sub> | 0.3 (-0.6 to 1.2) | 0.1 (-0.8 to 1.0) | 0.2 (-0.7 to 1.1) |
| D<sub>5</sub> | 0.4 (-0.5 to 1.3) | 0.4 (-0.5 to 1.2) | 0.0 (-0.9 to 0.9) |
| D<sub>6</sub> | 0.1 (-0.8 to 1.0) | -0.0 (-0.9 to 0.9) | 0.1 (-0.8 to 1.0) |
| D<sub>7</sub> | -0.2 (-1.1 to 0.7) | -0.2 (-1.1 to 0.7) | -0.0 (-0.9 to 0.9) |
| H + E, Heatwrap and exercise; H, Heatwrap; ODI, Oswestry Disability Index; NPRS, Numerical Pain Rating Scale; TSK-17, 17-item Tampa Scale of Kinesiophobia; CDSES-6, Chronic Disease Self-Efficacy Scale short version; PCS, Pain Catastrophizing Scale; GRoC, 15-point global rating of change.<br>*p<0.01 |  |  |  |

### Secondary outcomes

Estimated means for all outcomes at all time-points are presented in Supplementary Table 1. No Time x Group interaction was observed in secondary outcomes except for the global rating of change (p=0.023). Pairwise comparisons revealed that participants in the heatwrap alone group reported a smaller perception of change compared to participants in the heatwrap and exercise group (MD −2.6, 95% CI −4.1 to −1.0), and to those in the sham wrap group (−2.5, 95% CI −4.1 to −0.9) at W_1_. A main effect of time was observed at all time-points from baseline for all secondary outcomes (Table 2). The mean decrease in pain-related disability scores of all groups together reached clinical significance (i.e. within-group MD <-9% [MCID])^29^ at W_4_ (MD −12.6, 95% CI −14.9 to −10.3) and at all following time-points to W_52_ when compared to baseline scores. A similar pattern was observed for the average pain intensity scores of all groups, but clinical significance was reached at W_1_ (i.e. within-group MD <-2 [MCID]),^42^ with a MD of −2.2 (95% CI −2.5 to −1.7), then to all following time-points to W_52_ when compared to baseline scores (Figure 2B). The number of participants in each group who were using interventions specifically for their LBP at each follow-up except W_1_ are presented in Supplementary Table 3.

### Sensitivity analyses

The per-protocol analysis included 88 participants (n=29 for heatwrap and exercise, n=31 for heatwrap alone, and n=28 for sham heatwrap), and its results are presented in Supplementary Table 2. We obtained similar results for all outcomes as in the intention-to-treat analyses, with no Group x Time interaction (F=0.601; p=0.812) and a Time effect (F=43.220; p<0.001, MD −7.5%, 95% CI −9.4 to −5.6 at W_1_) for the primary outcome (ODI). The sensitivity analysis for ODI including exercises used as a co-intervention for the treatment of LBP as a covariate showed similar results as the primary analysis, with no Group x Time interaction (F=0.390; p=0.924) and a Time effect (F=50.746; p<0.001).

### Harms

Minor adverse events occurred in 7 participants. In the heatwrap and exercise group, one participant reported general discomfort caused by the heatwrap. In the heatwrap alone group, two participants reported a discomfort of the abdomen due to the tightness of the heatwrap, one reported a general discomfort with the heatwrap, and one reported redness, sweat and itching of the skin underneath the wrap. In the sham wrap group, one participant reported sweat and itching of the skin underneath the wrap, and one reported discomfort due to the wrap that was too tight. No harm related to exercise was reported. All participants who reported adverse events completed the intervention week.

## Discussion

The results of this trial showed that the combination of heatwrap and exercise was not more effective than either a heatwrap alone or a sham heatwrap to improve symptoms and functional outcomes on short-, mid- and long-term, when used in the acute phase of LBP, which is contrary to our initial hypothesis. Moreover, the results are opposed to those of prior trials on heatwrap in the treatment of acute and subacute LBP,^14–21^ and they do not support guideline recommendations for acute LBP.^10^

The strengths of this RCT include its independence from the heatwrap industry, which could partly explain the discrepancies between our results and those of prior studies.^14–21^ As previously mentioned, all studies using heatwrap in acute LBP were funded by the heatwrap manufacturer, and industry-sponsored studies were shown to be more likely to reach favorable efficacy outcomes compared to independent studies.^43^ A limitation arising from this is that we might have underestimated the needed sample size as we based its calculation on the potentially inflated effect sizes of the prior trials. However, between-group MDs in ODI scores at W_1_ in our study were all 1.6% and below, which is far from the MCID of 9%. It would thus be unlikely to find a clinically important difference with a larger participants’ sample. Another limitation is the potential influence of the exercises given to all participants after the intervention week on the mid- and long-term outcomes. However, when we adjusted our LMM with the use of exercises in our sensitivity analyses, the results remained similar. Also, we used a convenience sampling method which could have led to include individuals who might not have sought care if not enrolled in the study. This could impede on the external validity to care settings such as emergency. We did not have a group of exercise only, so we cannot definitively confirm that exercise was not effective against a sham heatwrap. However, as the heatwrap alone was not more effective than the sham heatwrap, it is unlikely that exercise alone would have provided a superior effect than that of exercise combined with a heatwrap. This is concordant with the recent Cochrane review on exercise in acute LBP that found evidence that exercise therapy was not better than no treatment or a placebo treatment to decrease pain and improve functional status^12^.

The discrepancy between our results and prior studies on heatwrap could also be explained by the fact that we provided all participants with evidence-based advice for LBP management and reassurance. We cannot confirm that the effect of advice was superior to the natural evolution of LBP, as we did not have a control group without advice. Our results were very similar to that of a systematic review and meta-analysis of Wallwork et al. (2024) on the natural course of LBP, which showed an important and rapid improvement in pain and disability levels in the first 6 weeks of acute LBP, followed by a slower and smaller improvement at 12 weeks, and stability in symptoms at 52 weeks.^2^ Nonetheless, advice is recommended as first-line care for patients with LBP.^5^ As most participants (57%) were categorised as low risk of developing persistent disability with the STarT Back Screening Tool,^26^ advice might have been enough to significantly decrease their disability and pain levels considering their good prognosis. Still, residual disability (7.3%, 95% CI 3.7 to 10.9) and pain (2.2/10, 95% CI 1.4 to 2.9) remained one year after the enrollment in the study. Future studies should examine if the effect of heatwrap and/or exercise differ in patients with acute LBP who are at moderate or high risk of persistent disability. Others could test a stepped-care approach as recommended in guidelines^5, 6^ by giving all participants with acute NSLBP advice and reassurance, then testing heatwrap and/or exercise with those who did not improve enough after 6 weeks.

Our results are concordant with the literature regarding the effect of exercise compared to a sham intervention or no intervention in acute LBP.^9, 12^ Nonetheless, considering the low number of trials on exercises in acute LBP, larger placebo-controlled trials should be conducted to confirm their effect in the treatment of acute LBP and strengthen recommendations on exercises in guidelines. Indeed, only two RCTs^44, 45^ assessed their effect against a placebo, and only two others^46, 47^ assessed their effect against no treatment in acute LBP, which limits the confidence in recommendations.

In conclusion, in this multi-arm RCT, the combination of heatwrap and exercise did not provide superior effect on symptoms and functional outcomes on short- mid and long-term when compared to a heatwrap alone and a sham heatwrap in acute LBP. We believe future guidelines should consider these results to formulate recommendations for acute LBP.

## Data Availability

All data produced in the present study are available upon reasonable request to the authors

