## Supplementary material for "Effects of heatwrap and exercise in acute low back pain: a multi-arm randomised controlled trial"

**Supplementary Table 1.** Outcomes for each group at all time-points presented as mean (95% CI)

|  | **Heatwrap and exercise** | **Heatwrap alone** | **Sham heatwrap** |
| --- | --- | --- | --- |
| ODI (0 to 100) | | | |
| W_0_ | 23.0 (19.8 to 26.2) | 22.8 (19.6 to 26.1) | 22.4 (19.1 to 25.6) |
| W_1_ | 14.2 (11.0 to 17.5) | 15.9 (12.6 to 19.1) | 14.7 (11.4 to 18.0) |
| W_4_ | 9.5 (6.2 to 12.8) | 10.0 (6.8 to 13.3) | 10.8 (7.4 to 14.2) |
| W_12_ | 8.3 (5.1 to 11.6) | 7.0 (3.6 to 10.4) | 8.9 (5.5 to 12.2) |
| W_26_ | 7.2 (3.8 to 10.6) | 8.9 (5.5 to 12.2) | 8.7 (5.3 to 12.2) |
| W_52_ | 7.3 (3.7 to 10.9) | 6.0 (2.7 to 9.3) | 5.3 (1.8 to 8.8) |
| NPRS (0 to 10) – Average pain intensity in the last 7 days | | | |
| W_0_ | 4.7 (4.1 to 5.4) | 5.4 (4.7 to 6.1) | 5.2 (4.5 to 5.9) |
| W_1_ | 3.1 (2.4 to 3.7) | 3.1 (2.4 to 3.8) | 2.7 (2.0 to 3.4) |
| W_4_ | 2.4 (1.7 to 3.1) | 2.5 (1.8 to 3.2) | 2.1 (1.4 to 2.8) |
| W_12_ | 1.7 (1.0 to 2.4) | 1.6 (0.9 to 2.3) | 2.6 (1.9 to 3.3) |
| W_26_ | 2.2 (1.5 to 2.9) | 1.7 (1.0 to 2.4) | 1.9 (1.2 to 2.7) |
| W_52_ | 2.2 (1.4 to 2.9) | 1.9 (1.2 to 2.5) | 1.7 (1.0 to 2.5) |
| TSK-17 (17 to 68) | | | |
| W_0_ | 36.8 (34.5 to 39.2) | 35.5 (33.1 to 37.9) | 36.7 (34.3 to 39.1) |
| W_1_ | 33.1 (30.7 to 35.4) | 33.3 (30.9 to 35.7) | 33.3 (30.9 to 35.7) |
| W_4_ | 31.6 (29.2 to 34.0) | 31.6 (29.2 to 33.9) | 32.3 (29.8 to 34.7) |
| W_12_ | 30.8 (28.4 to 33.2) | 32.2 (29.8 to 34.6) | 31.3 (28.9 to 33.8) |
| W_26_ | 32.6 (30.2 to 35.1) | 31.5 (29.1 to 33.9) | 32.2 (29.7 to 34.7) |
| W_52_ | 31.6 (29.1 to 34.2) | 30.8 (28.3 to 33.2) | 31.5 (28.9 to 34.0) |
| CPSES-6 (6 to 60) | | | |
| W_0_ | 7.8 (7.2 to 8.3) | 7.8 (7.3 to 8.3) | 8.0 (7.5 to 8.5) |
| W_1_ | 8.2 (7.7 to 8.8) | 8.3 (7.8 to 8.8) | 8.2 (7.7 to 8.8) |
| W_4_ | 8.7 (8.1 to 9.2) | 8.5 (7.9 to 9.0) | 8.6 (8.1 to 9.2) |
| W_12_ | 8.7 (8.2 to 9.2) | 8.5 (8.0 to 9.1) | 8.7 (8.1 to 9.2) |
| W_26_ | 8.5 (7.9 to 9.0) | 8.6 (8.0 to 9.1) | 8.3 (7.7 to 8.8) |
| W_52_ | 8.4 (7.9 to 9.0) | 8.8 (8.3 to 9.4) | 8.8 (8.2 to 9.4) |
| PCS (0 to 52) | | | |
| W_0_ | 12.9 (10.1 to 15.7) | 16.4 (13.6 to 19.2) | 14.3 (11.5 to 17.2) |
| W_1_ | 8.7 (5.9 to 11.5) | 10.8 (7.9 to 13.7) | 8.7 (5.8 to 11.6) |
| W_4_ | 4.4 (1.5 to 7.3) | 8.5 (5.7 to 11.4) | 7.7 (4.7 to 10.6) |
| W_12_ | 3.8 (0.9 to 6.7) | 7.9 (5.0 to 10.9) | 6.3 (3.3 to 9.2) |
| W_26_ | 3.7 (0.7 to 6.6) | 7.2 (4.3 to 10.1) | 5.6 (2.6 to 8.6) |
| W_52_ | 6.4 (3.3 to 9.5) | 8.7 (5.7 to 11.7) | 5.0 (1.9 to 8.2) |
| GRoC (-7 to 7) | | | |
| W_1_ | 3.9 (2.8 to 4.9) | 1.3 (0.2 to 2.4) | 3.7 (2.6 to 4.9) |
| W_4_ | 4.2 (3.1 to 5.3) | 3.7 (2.5 to 4.8) | 4.2 (3.1 to 5.4) |
| W_12_ | 4.3 (3.2 to 5.4) | 4.4 (3.3 to 5.5) | 3.5 (2.3 to 4.6) |
| W_26_ | 4.0 (2.9 to 5.2) | 4.8 (3.7 to 5.9) | 4.2 (3.0 to 5.3) |
| W_52_ | 4.2 (3.0 to 5.4) | 4.3 (3.2 to 5.4) | 4.0 (2.7 to 5.2) |
| NPRS (0 to 10) – Average daily pain intensity | | | |
| D_1_ | 3.8 (3.1 to 4.4) | 3.3 (2.7 to 3.9) | 3.0 (2.4 to 3.7) |
| D_2_ | 3.3 (2.6 to 3.9) | 2.9 (2.3 to 3.5) | 2.4 (1.8 to 3.1) |
| D_3_ | 2.9 (2.3 to 3.5) | 2.6 (2.0 to 3.2) | 2.3 (1.6 to 2.9) |
| D_4_ | 2.8 (2.2 to 3.4) | 2.6 (1.9 to 3.2) | 2.5 (1.8 to 3.1) |
| D_5_ | 2.7 (2.1 to 3.3) | 2.7 (2.1 to 3.3) | 2.3 (1.7 to 3.0) |
| D_6_ | 2.6 (2.0 to 3.2) | 2.5 (1.9 to 3.1) | 2.5 (1.9 to 3.1) |
| D_7_ | 2.1 (1.4 to 2.7) | 2.1 (1.5 to 2.8) | 2.3 (1.6 to 2.9) |
| ODI, Oswestry Disability Index; NPRS, Numerical Pain Rating Scale; TSK-17, 17-item Tampa Scale of Kinesiophobia; CPSES-6, Chronic Pain Self-Efficacy Scale short version; PCS, Pain Catastrophizing Scale; GRoC, 15-point global rating of change; W_0_, baseline; W_1_, week 1; W_4_, week _4_; W_12_, week 12; W_26_, week 26; W_52_, week 52; D_1_, day 1; D_2_, day 2; D_3_, day 3; D_4_, day 4; D_5_, day 5; D_6_, day 6; D_7_, day 7. | | | |

**Supplementary Table 2.** Per-protocol analysis

| **ODI** | |
| --- | --- |
| Group x Time | F=0.601; p=0.812 |
| Time | F=43.220; p<0.001 |
| **NPRS*** | |
| Group x Time | F=1.763; p=0.068 |
| Time | F=42.758; p<0.001 |
| **TSK-17** | |
| Group x Time | F=0.345; p=0.968 |
| Time | 16.430; p<0.001 |
| **CPSES-6** | |
| Group x Time | F=0.796; p=0.633 |
| Time | F=5.276; p<0.001 |
| **PCS** | |
| Group x Time | F=0.448; p=0.921 |
| Time | F=21.462; p<0.001 |
| **GRoC** | |
| Group x Time | F=2.441; p=0.015^a^ |
| Time | F=3.207; p=0.014 |
| **NPRS daily**** | |
| Group x Time | F=0.753; p=0.699 |
| Time | F=9.576; p<0.001 |
| ODI, Oswestry Disability Index; NPRS, Numerical Pain Rating Scale; TSK-17, 17-item Tampa Scale of Kinesiophobia; CPSES-6, Chronic Pain Self-Efficacy Scale short version; PCS, Pain Catastrophizing Scale; GRoC, 15-point global rating of change.  *****Average pain intensity in the last 7 days.  **Average daily pain intensity.  ^a^Pairwise comparisons: heatwrap and exercise (MD 2.3, 95% CI 0.639 to 3.893; p=0.007) and sham heatwrap (MD 2.9, 95% CI 1.272 to 4.550; p<0.001) groups vs heatwrap alone. | |

**Supplementary Table 3**. Percentage of participants in each group using co-interventions for their LBP at baseline and at 4-, 12-, 26- and 52-weeks follow-ups

|  | **Heatwrap and exercise (n=34)** | **Heatwrap alone (n=33)** | **Sham heatwrap (n=32)** |
| --- | --- | --- | --- |
| Exercises | | | |
| W_0_ | 15 | 39 | 28 |
| W_4_ | 45 | 50 | 76 |
| W_12_ | 44 | 50 | 58 |
| W_26_ | 28 | 37 | 39 |
| W_52_ | 31 | 44 | 44 |
| Opioids | | | |
| W_0_ | 0 | 0 | 3 |
| W_4_ | 3 | 10 | 7 |
| W_12_ | 6 | 0 | 0 |
| W_26_ | 0 | 0 | 0 |
| W_52_ | 4 | 0 | 0 |
| Corticosteroid injection | | | |
| W_0_ | 0 | 0 | 0 |
| W_4_ | 0 | 3 | 0 |
| W_12_ | 6 | 0 | 3 |
| W_26_ | 4 | 0 | 4 |
| W_52_ | 4 | 0 | 0 |
| Psychological support | | | |
| W_0_ | 0 | 3 | 0 |
| W_4_ | 3 | 3 | 0 |
| W_12_ | 9 | 0 | 0 |
| W_26_ | 0 | 0 | 0 |
| W_52_ | 4 | 0 | 0 |
| W_0_, baseline; W_4_, week _4_; W_12_, week 12; W_26_, week 26; W_52_, week 52 | | | |
